# Identifying novel factors associated with COVID-19 transmission and fatality using the machine learning approach

**DOI:** 10.1101/2020.06.10.20127472

**Authors:** Mengyuan Li, Zhilan Zhang, Wenxiu Cao, Yijing Liu, Beibei Du, Canping Chen, Qian Liu, Md. Nazim Uddin, Shanmei Jiang, Cai Chen, Yue Zhang, Xiaosheng Wang

## Abstract

The COVID-19 virus has infected millions of people and resulted in hundreds of thousands of deaths worldwide. By using the logistic regression model, we identified novel critical factors associated with COVID19 cases, death, and case fatality rates in 154 countries and in the 50 U.S. states. Among numerous factors associated with COVID-19 risk, we found that the unitary state system was counter-intuitively positively associated with increased COVID-19 cases and deaths. Blood type B was a protective factor for COVID-19 risk, while blood type A was a risk factor. The prevalence of HIV, influenza and pneumonia, and chronic lower respiratory diseases was associated with reduced COVID-19 risk. Obesity and the condition of unimproved water sources were associated with increased COVID-19 risk. Other factors included temperature, humidity, social distancing, smoking, and vitamin D intake. Our comprehensive identification of the factors affecting COVID-19 transmission and fatality may provide new insights into the COVID-19 pandemic and advise effective strategies for preventing and migrating COVID-19 spread.

## Introduction

Since its first report in December 2019, COVID-19 caused by the 2019 novel coronavirus (SARS-CoV-2) has spread across 215 countries and territories ^1^. As of May 15, 2020, more than 4,500,000 COVID-19 cases and 300,000 deaths were reported ^1^. Compared to other coronaviruses, such as SARS-CoV and MERS-CoV, SARS-CoV-2 has a significantly higher infectivity potential that makes it spread across the world rapidly and has caused a global pandemic ^2^. Meanwhile, the fatality rate for COVID-19 is not low, making SARS-CoV-2 more destructive than the previously emerging coronaviruses ^3^. The COVID-19 pandemic has established the most critical global health and economic crisis in recent years ^4^. But even worse, its development is difficult to forecast ^5^. Previous studies have identified individual factors associated with the spread of COVID-19. For example, temperature and humidity may impact the transmission of COVID-19 ^6 7^. Social distancing can significantly reduce COVID-19 transmission ^8 9^. Besides, some factors associated with the COVID-19 death risk have been identified, such as age, sex, and comorbidities ^10 11^. Some factors, such as diet and nutrition ^12 13^, have been associated with both COVID-19 infection and mortality. Nevertheless, one standard limitation of these studies is that they ignored the interdependence between different factors. As a result, they might not accurately describe the relationships between some factors and COVID-19.

To simultaneously identify different factors affecting COVID-19 transmission and fatality, we used logistic models to predict population-adjusted confirmed cases (per one million) and deaths (per one million) and case fatality rates (CFRs) of COVID-19 in different countries and in the 50 U.S. states. We collected data related to politics, economy, culture, demographic, geography, education, medical resource, scientific development, environment, diseases, diet, and nutrition. Based on these data, we defined tens of factors (variables) that could affect COVID-19 transmission and fatality. We used least absolute shrinkage and selection operator (LASSO) ^14^ to select the factors associated with COVID-19 risk. We identified novel factors associated with COVID-19 risk. We also confirmed some controversial factors and most of the well-recognized factors associated with COVID-19 risk.

## Results

### Identifying factors affecting COVID-19 transmission and fatality in different countries

We first used the LASSO to evaluate the contribution of the 77 variables to COVID-19 cases, deaths, or CFRs. The Lasso selected 57 variables with a nonzero β-coefficient in at least one of the three predictive models for COVID-19 cases, deaths, and CFRs (Fig. 1). As expected, the three social distancing-associated factors (large-scale sports, religious, and cultural events) represented positive predictors of COVID-19 deaths and/or CFRs. The Lasso selected two temperature types (medium and low) in at least one predictive model. The medium temperature was a positive predictor of COVID-19 deaths, while the low temperature was a negative predictor of COVID-19 deaths and CFRs. These data, together with the findings from previous studies ^6 7^, suggest that the risk of COVID-19 transmission is high in a preferential temperature range, but it will decline with the temperature rising or falling outside the range. The humidity was a negative predictor of COVID-19 cases, consistent with the finding from a previous study ^7^. The negative association between humidity and COVID-19 risk was following two other findings: i) the island country was a negative predictor of COVID-19 deaths (the weather is humid in island countries), and ii) the arid climate was a positive predictor of COVID-19 deaths. Besides, we found that the temperate climate was a positive predictor of COVID-19 CFRs, while the cold climate was a negative predictor of COVID-19 CFRs. Again, these results suggest that temperature has a prominent association with COVID-19 risk. Surprisingly, the cold climate was a positive predictor of COVID-19 cases, in contrast with its negative association with CFRs. The latitude was a negative predictor of COVID-19 deaths and CFRs, mainly because it is one of the main factors that affect temperature, which has an essential association with COVID-19 risk. In contrast, the longitude was a positive predictor of COVID-19 cases and deaths. The sex ratio (male/female) was a positive predictor of COVID-19 cases, while the percentage of the population aged < 10 years was a negative predictor of COVID-19 cases and deaths. It is in agreement with previous findings that sex and age were significantly associated with COVID-19 risk ^10 11^. Islam was a positive predictor of COVID-19 cases, and Christianism was a positive predictor of CFRs. In contrast, Buddhism and other religions were negative predictors of COVID-19 cases, deaths, and/or CFRs. These results could be associated with the fact that Islam and Christianism have relatively large and regular gathering activities that facilitate the transmission of COVID-19. Among the four races, the White was a positive predictor of COVID-19 deaths, and the Asian and Arab were negative predictors of COVID-19 cases. These results conform to the fact that the countries reporting a large number of COVID-19 deaths were mainly majority-white countries. In fact, among the 24 countries with COVID-19 deaths > 1,000 as of May 15, 2020, 18 countries were majority white, including 14 European and 4 American countries (USA, Canada, Brazil, and Mexico). Interestingly, we found that particular food or nutrition intake was associated with COVID-19 risk. For example, increased intake of vegetables, grain, sugar, protein, fat, vitamin D, and vitamin K was associated with reduced COVID-19 cases, deaths, and/or CFRs. In contrast, increased intake of fruits, dairy products, vitamin B, and vitamin C was associated with increased COVID-19 cases, deaths, and/or CFRs. The negative association between vitamin D intake and COVID-19 risk is consistent with previous reports ^12 13^. The condition of unimproved water sources was a positive predictor of CFRs, suggesting the potential of COVID-19 spreading through water. The air passenger traffic and vehicle usage were a positive and a negative predictor of COVID-19 deaths, respectively, and the railway length was not selected by the Lasso for predicting COVID-19 cases, deaths, or CFRs. This indicates that air transport is the most dangerous approach for COVID-19 transmission compared to other modes of transport. It is reasonable since small closed spaces on airplanes are prone to virus transmission. The percentage of urban population was a positive and a negative predictor of COVID-19 cases and CFRs, respectively. A possible explanation is that dense urban population is conducive to COVID-19 transmission, while excellent urban medical conditions are conducive to the treatment of COVID-19 patients. The average age of childbirth was a positive predictor of COVID-19 cases, deaths, and CFRs, and the life expectancy was a positive predictor of COVID-19 deaths. These results suggest a positive association between age and COVID-19 risk.

**Fig. 1.**
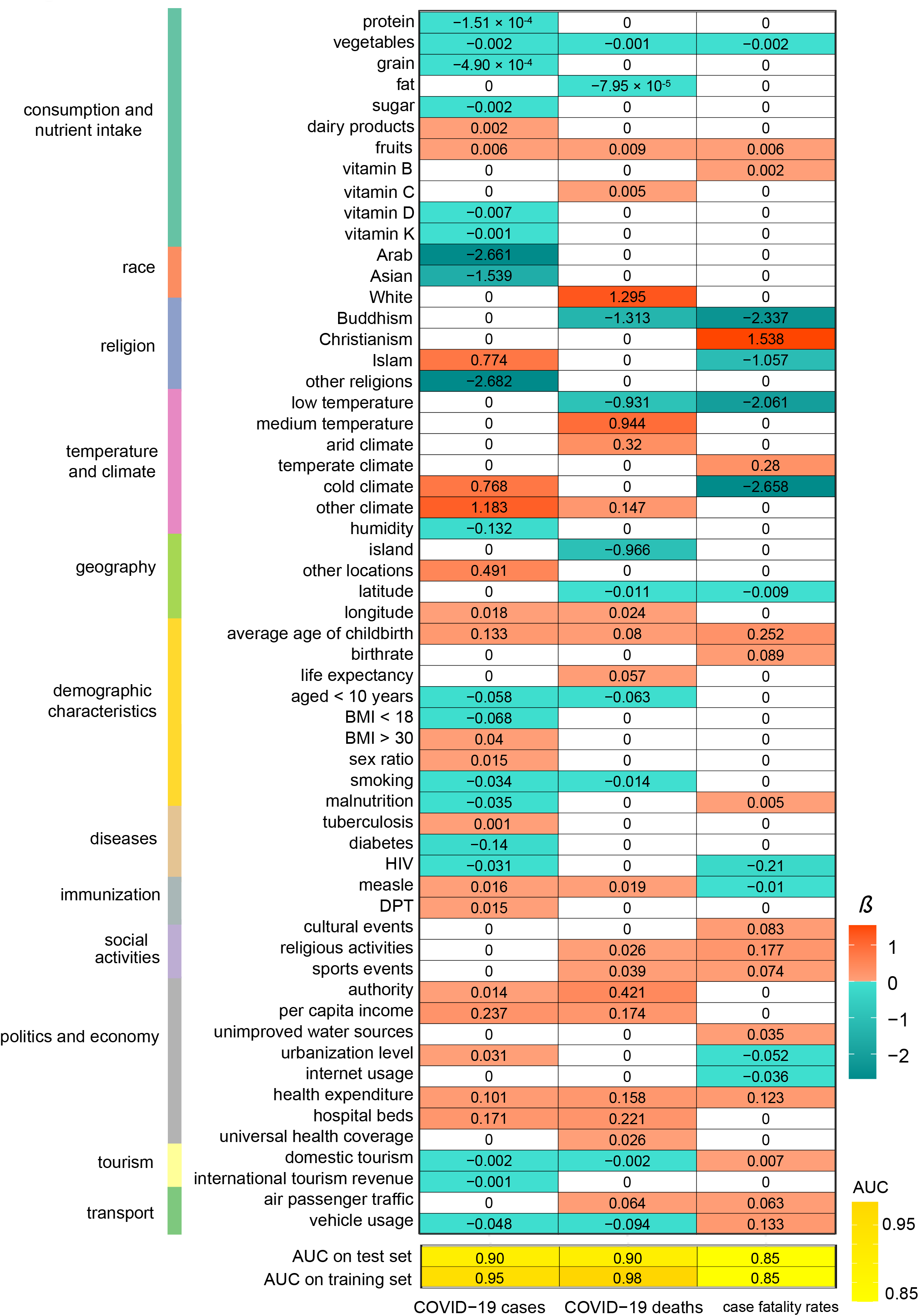
57 variables selected by the LASSO in predicting COVID-19 cases, deaths, and/or case fatality rates in 154 countries. The *β*-coefficients and AUCs in the ridge logistic models are shown. AUC: the area under the receiver operating characteristic curve; aged < 10 years: percentage of population aged < 10 years; Arab: percentage of the Arab population; Asian: percentage of the Asian population; authority: governing system (unitary state or federation); BMI < 18: percentage of population with body mass index (BMI) < 18; BMI > 30: percentage of population with body mass index (BMI) > 30; Buddhism: Buddhism as the primary religion; Christianism: Christianism as the primary religion; cold climate: cold (continental) climate; cultural events: number of participants in large-scale cultural events; dairy products: dairy product intake (kcal/capita/day); diabetes: prevalence of diabetes; domestic tourism: domestic tourism expenditure (billion dollars); DPT: DPT (diphtheria, pertussis, tetanus) immunization coverages among children ages 12-23 months; fat: fat intake (kcal/capita/day); fruits: fruit intake (kcal/capita/day); grain: grain intake (kcal/capita/day); health expenditure: government expenditure on health per capita; HIV: prevalence of HIV; hospital beds: hospital beds per 1,000 people; humidity: relative humidity; internet usage: percentage of population using the internet; Islam: Islam as the primary religion; island: geographic location-island country; malnutrition: percentage of population of malnutrition; measle: measle immunization coverages among children ages 12-23 months; other locations: geographic location-other country; other religions: other religions as the primary religion or without religion; protein: protein intake (kcal/capita/day); religious activities: number of participants in large-scale religious activities; sex ratio: sex ratio (number of males per 100 females); smoking: smoking rate among people aged more than 15 years old; sports events: number of participants in major sports events; sugar: sugar intake (kcal/capita/day); tuberculosis: prevalence of tuberculosis; unimproved water sources: percentage of population using unimproved water sources; urbanization level: percentage of urban population; vegetables: vegetable intake (kcal/capita/day); vehicle usage: number of vehicles; vitamin B: vitamin B (B6, B9, and B12) intake (kcal/capita/day); vitamin C: vitamin C intake (kcal/capita/day); vitamin D: vitamin D intake (kcal/capita/day); vitamin K: vitamin K intake (kcal/capita/day); White: percentage of the White population.

We also obtained some unexpected findings. For example, the economic development (per capita income) and health investment (universal health coverage index, health expenditure, and hospital beds per 1,000 people) were positive predictors of COVID-19 cases, deaths, and/or CFRs. The smoking rate was a negative predictor of COVID-19 cases and deaths, consistent with the recent finding that smoking was a protective factor for COVID-19 infection ^15^. Nevertheless, an association between smoking and COVID-19 CFRs was not observed in our model, suggesting that smoking is not likely to reduce the mortality risk of COVID-19 patients. The prevalence of some diseases, such as HIV and diabetes, were inversely associated with COVID-19 risk. The prevalence of malnutrition was inversely associated with COVID-19 cases but was positively associated with COVID-19 CFRs. The percentages of the population with BMI < 18 and BMI > 30 were negatively and positively associated with COVID-19 cases, respectively, suggesting that obesity could be a risk factor for COVID-19. The unitary state system was a positive predictor of COVID-19 cases and deaths, suggesting that it is less effective in controlling COVID-19 outbreaks than the federation system. This conflicts with common sense that the centralization system is capable of concentrating resources more efficiently than the decentralization system in fighting against COVID-19.

### Identifying factors affecting COVID-19 transmission and fatality in the 50 U.**S. states**

To uncover the critical factors for COVID-19 risk within a single country, we used logistic models to predict COVID-19 cases, deaths, and CFRs in the 50 U.S. states. We used 55 variables in the predictive models, most of which were in the list of variables mentioned earlier. Likewise, we first used the LASSO to determine the contribution of the 55 variables to COVID-19 cases, deaths, or CFRs. We defined the states as having low or high COVID-19 cases, deaths, or CFRs based on their median values. The next steps followed the previous method. The Lasso selected 31 variables with a nonzero *β*-coefficient in at least one of the three predictive models for COVID-19 cases, deaths, and CFRs (Fig. 2). Consistent with the previous results, medium temperature, arid climate, social distancing (major sports events), per capita income, longitude, and the average age of childbirth were positive predictors of COVID-19 cases, deaths, and/or CFRs, and humidity, smoking rate, and international tourism revenue were negative predictors. The urbanization level was a positive predictor of COVID-19 cases, while it was a negative predictor of COVID-19 deaths and CFRs. We have also obtained some findings that were special in the U.S. setting. For example, the death rate of influenza and pneumonia, and chronic lower respiratory diseases were negative predictors of COVID-19 cases, deaths, and/or CFRs. A potential explanation of the negative association between the prevalence of influenza and pneumonia and COVID-19 risk is that there is cross-reactive immunity between other types of viruses and SARS-CoV-2 ^16^. The prevalence of diabetes was a positive predictor of COVID-19 CFRs, indicating that it is a risk factor for COVID-19 fatality. However, in both world and U.S. settings, it was a negative predictor of COVID-19 cases, indicating that it is a protective factor for COVID-19 infection. Health insurance coverage was positively associated with COVID-19 cases, deaths, and CFRs. The level of CO2 emissions per capita was negatively associated with COVID-19 cases and deaths. The Black was a positive predictor of COVID-19 cases and deaths, consistent with previous reports ^17^. The geographic location correlated with COVID-19 risk where landlocked country and coastal region had lower risk and the great lakes region had a higher risk. Besides, the usage of private transport in urban might reduce COVID-19 risk.

**Fig. 2.**
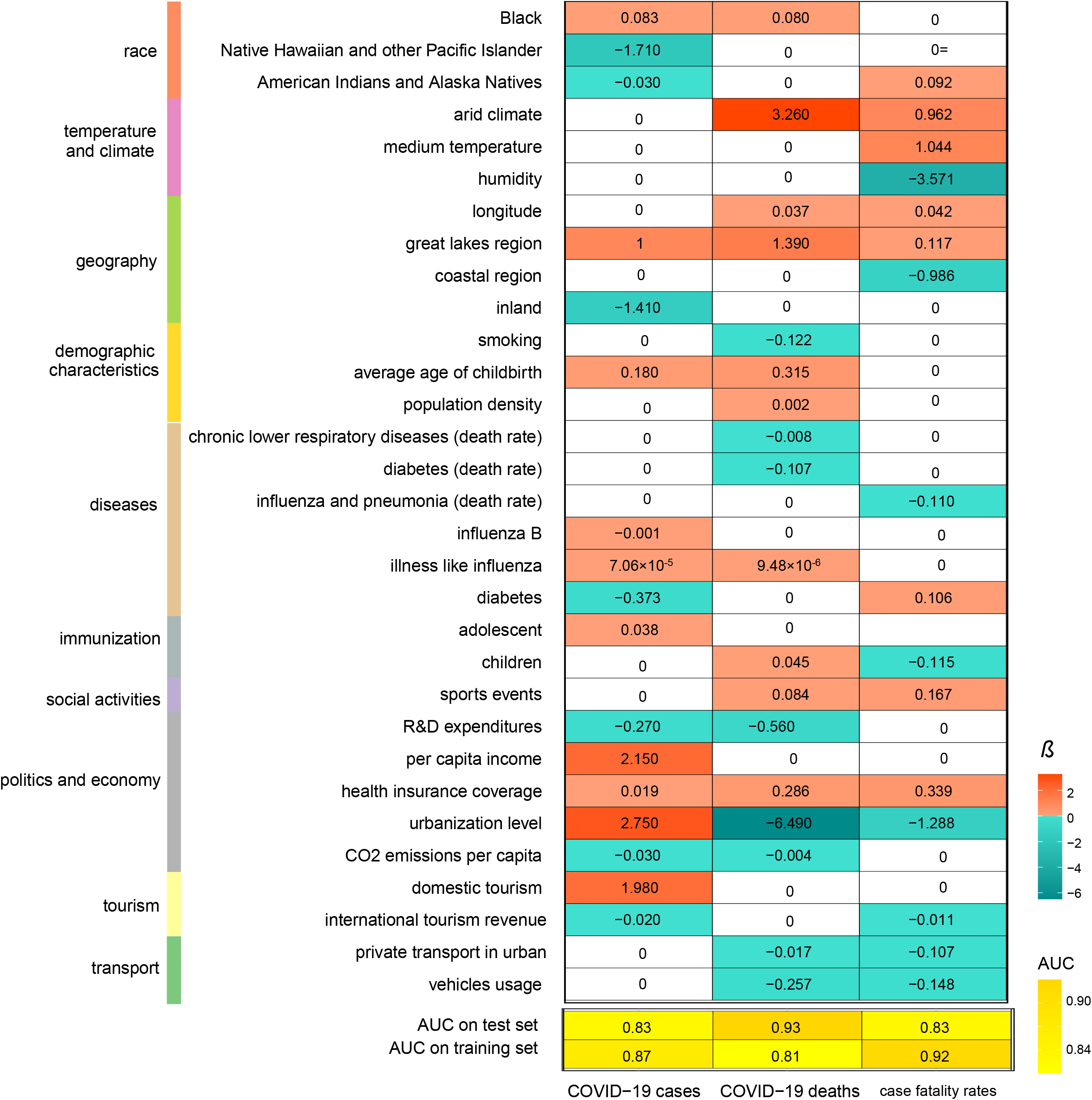
31 variables selected by the LASSO in predicting COVID-19 cases, deaths, and/or case fatality rates in the U.S. 50 states. The *β*-coefficients and AUCs in the ridge logistic models are shown. adolescent: percentage of adolescents 11 - 17 years with 2+ Adolescent Immunizations; American Indians and Alaska Natives: percentage of the American Indians and Alaska Natives population; Black: percentage of the Black population; children: percentage of children < 6 years with 2+ Immunizations; chronic lower respiratory diseases (death rate): deaths from chronic lower respiratory diseases per 100,000 population; coastal region: geographic location-coastal region; diabetes: prevalence of diabetes; diabetes (death rate): deaths from diabetes per 100,000 population; domestic tourism: mean census estimate of vehicle trips in urban; great lakes region: geographic location-great lakes region; health insurance coverage: percentage of health insurance coverage; humidity: relative humidity; influenza and pneumonia (death rate): deaths from influenza and pneumonia per 100,000 population; influenza B: number of positive influenza B (Victoria and Yamagata Lineage); influenza-like illness: number of patients with influenza-like illness; inland: geographic location-landlocked country; international tourism revenue: travel spending by international visitors in the U.S. (billion); Native Hawaiian and other Pacific Islander: percentage of the White population; per capita income: per capita real income; private transport in urban: mean census estimate of vehicle miles traveled in urban; smoking: percentage of smoking population; sports events: number of live audiences in major sports events; urbanization level: percentage of urban population; vehicle usage: proportion of households with cars.

### Ranking variables based on their importance in distinguishing between low and high COVID-19 cases, deaths, and CFRs

In predicting COVID-19 cases, deaths, and CFRs, we achieved the AUC values of 0.90, 0.90, and 0.85, respectively, on the training set, and 0.95, 0.98, and 0.85, respectively, on the test set in countries, and all AUC values > 0.8 in the 50 U.S. states (Figs. 1 and 2). These results demonstrate that the variables selected by the LASSO represent important factors affecting COVID-19 risk. We ranked the importance of variables in distinguishing between low and high COVID-19 cases, deaths, and CFRs based on the chi-square statistic. In the world setting, per capita income, malnutrition, aged < 10 years, urbanization level, dairy product intake, BMI < 18, hospital beds, BMI > 30, longitude, and protein intake were the ten most essential variables in distinguishing between low and high COVID-19 cases (Fig. 3A). The ten most important variables in distinguishing between low and high COVID-19 deaths were life expectancy, aged < 10 years, universal health coverage, per capita income, longitude, vitamin C intake, fat intake, hospital beds, domestic tourism, and health expenditure (Fig. 3A). Latitude, measle immunization, major sports events, internet usage, urbanization level, malnutrition, unimproved water sources, vehicle usage, cold climate, and domestic tourism were the ten most important variables in distinguishing between low and high COVID-19 CFRs (Fig. 3A). These results collectively demonstrate that age, climate, social distancing, economic development, health investment, weight, and nutrient intake are mostly associated with COVID-19 risk. We obtained similar results in the U.S. setting except influenza and race being two prominent factors for COVID-19 risk (Fig. 3B).

**Fig. 3.**
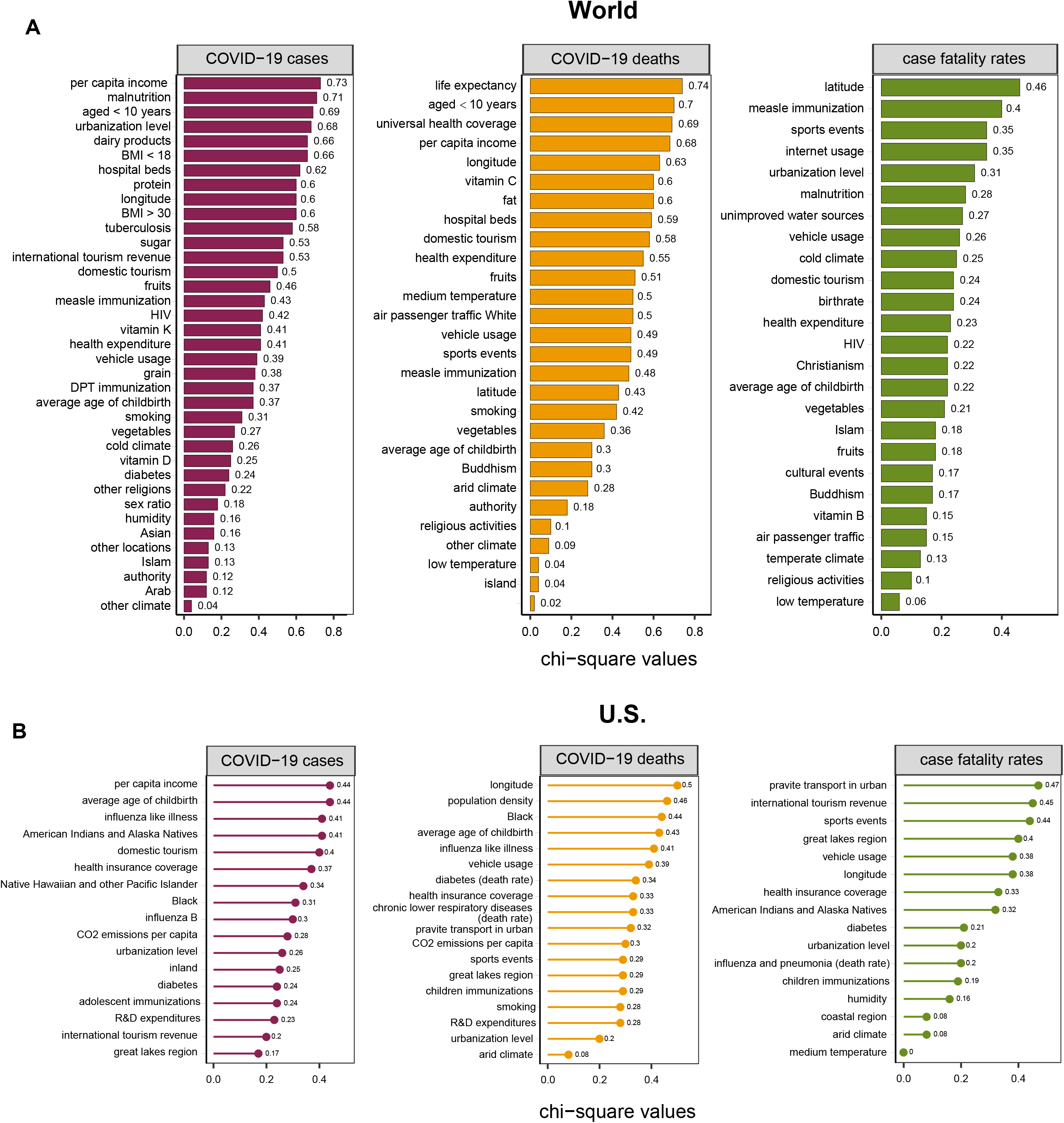
Ranking the importance of variables in distinguishing between low and high COVID-19 cases, deaths, and CFRs based on the chi-square statistic. The importance of variables in the world **(A)** and the U.S. **(B)** settings. These variables were selected by the LASSO. The chi-square values are shown.

### Associations of blood type, social distancing, and temperature with COVID-19 transmission and fatality

To investigate the association between the ABO blood type and COVID-19 vulnerability, we also collected ABO blood type distribution (population proportion) by country. We obtained the complete ABO blood type distribution data in 94 countries. We found that blood type A had significant positive correlations with COVID-19 cases (Spearman’s correlation test, *ρ* = 0.47, *P* = 1.97 × 10^−6^) and deaths (*ρ* = 0.58, *P* = 6.23 × 10^−10^) (Fig. 4A). The correlation between blood type A and COVID-19 CFRs was not significant (*ρ* = 0.18, *P* = 0.09). Overall, these results indicate that blood type A is associated with an increased risk of COVID-19, consistent with a recent report ^18^. In contrast, blood type B displayed significant inverse correlations with COVID-19 cases (*ρ* = -0.41, *P* = 3.91 × 10^−5^), deaths (*ρ* = -0.50, *P* = 2.63 × 10^−7^), and CFRs (*ρ* = -0.34, *P* = 8.91 × 10^−4^) (Fig. 4A). These results indicate that blood type B is associated with a reduced risk of COVID-19. The association between the ABO blood type and COVID-19 vulnerability may partially explain why the COVID-19 outbreak is serious in Europe and relatively moderate in South Asia since blood type A has a high frequency in Europe, and blood type B has a high frequency in South Asia ^19 20^.

**Fig. 4.**
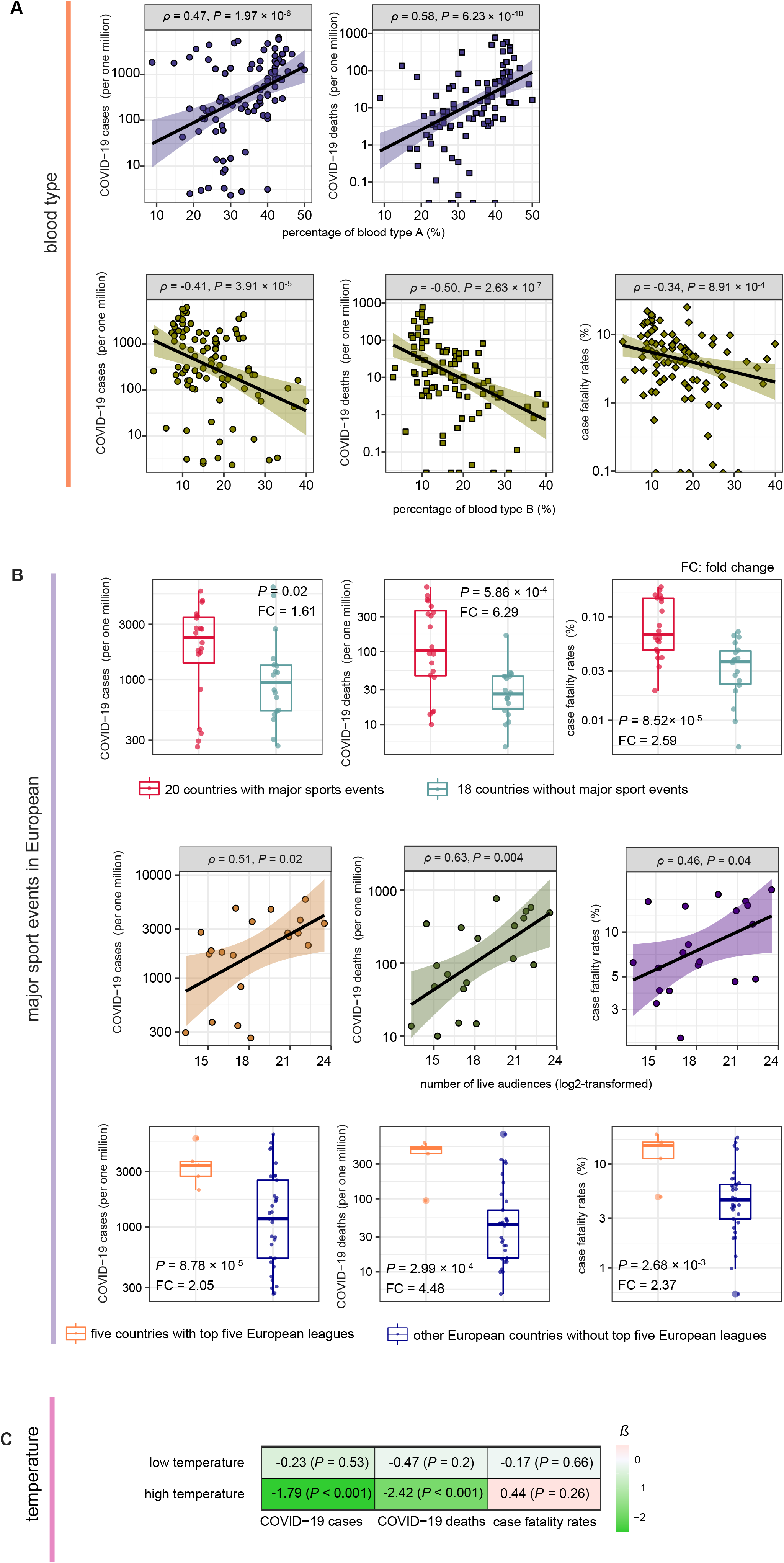
Associations of the ABO blood type, major sports events, and temperature with COVID-19. **(A)** Blood type A is a risk factor for COVID-19, and blood type B is a protective factor. **(B)** The positive association between major sports events and COVID-19 risk in European countries. **(C)** High temperature has a significantly stronger power than low temperature in predicting COVID-19 cases and deaths. The *β*-coefficients and *P* values in logistic regression models are shown.

To investigate the association between social distancing and COVID-19 spread, we analyzed the correlations between major sports events from January 2020 to March 2020 and COVID-19 cases, deaths, and CFRs in 38 European countries. We focused on European countries because sports, particularly football, are highly popular in most countries in Europe. We classified 38 European countries into two groups. The first group contained 20 countries hosting major sports events, including Six Nations Championship, UEFA Europa League, and domestic football matches from January 2020 to March 2020, and the second group contained 18 countries hosting no major sports events during this period. We found that COVID-19 cases, deaths, and CFRs were significantly higher in the first group than in the second group (one-sided Mann– Whitney U test, *P* < 0.01) (Fig. 4B). The mean COVID-19 cases, deaths, and CFRs were 1.61, 6.29, and 2.59 times higher in the first group than that in the second group, respectively. Moreover, within the 20 countries hosting major sports events, the total numbers of participants in the major sports events had significant positive correlations with COVID-19 cases (*ρ* = 0.51, *P* = 0.02), deaths (*ρ* = 0.63, *P* = 0.004), and CFRs (*ρ* = 0.46, *P* = 0.04) (Fig. 4B). Football is the most popular sport in almost all European countries, and the top five European leagues were subject to a large number of live audiences. As expected, the five countries with the top five European leagues, including UK, Spain, Italy, France, and Germany, had significantly higher COVID-19 cases, deaths, and CFRs than the other European countries (one-sided Mann–Whitney U test, *P* < 0.01, fold change > 2) (Fig. 4B). Altogether, these results suggest that major sports events can boost COVID-19 transmission and highlight the importance of social distancing in mitigating COVID-19 spread.

The association between temperature and COVID-19 transmission has been well recognized ^6 7 21^. To compare the relative contribution of high and low temperature in predicting COVID-19 risk, we built logistic models with both variables to predict COVID-19 cases, deaths, and CFRs in the 154 countries. As expected, both high and low temperatures were negative predictors of COVID-19 cases and deaths. However, the high temperature displayed a significantly stronger power than the low temperature in predicting COVID-19 cases and deaths (Fig. 4C). This indicates that high temperature is a more compelling factor mitigating COVID-19 transmission than low temperature.

## Discussion

We comprehensively described the most associated factors affecting COVID-19 transmission and fatality. Among numerous factors associated with COVID-19 risk, the unitary state system was counter-intuitively positively associated with increased COVID-19 cases and deaths. Blood type B was a protective factor for COVID-19 risk, while blood type A was a risk factor. The prevalence of HIV, influenza and pneumonia, and chronic lower respiratory diseases was associated with reduced COVID-19 risk. Obesity and the condition of unimproved water sources were associated with increased COVID-19 risk. Other factors included temperature, humidity, social distancing, smoking, and vitamin D intake. The factors most associated with COVID-19 risk included age, climate, social distancing, economic development, health investment, weight, and nutrient intake.

This study is interesting for several reasons. Firstly, we used the LASSO to evaluate the contribution of different variables to COVID-19 cases, deaths, or case fatality rates (CFRs) and identified the most critical factors associated with COVID-19 based on data related to politics, economy, culture, demographic, geography, education, medical resource, scientific development, environment, diseases, diet, and nutrition. This method overcame the limitation of previous univariate analyses that ignored the interdependence between different factors. Second, we identified novel factors associated with COVID19, including the unitary state governing system as a positive predictor of COVID-19 cases and deaths, blood type B as a protective factor for COVID-19 risk, the negative associations between the prevalence of HIV, influenza and pneumonia, and chronic lower respiratory diseases and COVID-19 risk, and the positive associations between economic development, education expenditure, obesity, and condition of unimproved water sources and COVID-19 risk. Third, we confirmed some controversial factors associated with COVID-19, including smoking and vitamin D intake as protective factors and blood type A as a risk factor for COVID-19. Finally, this study demonstrates that age, climate, social distancing, economic development, health investment, weight, nutrient intake, influenza, and race are the most prominent factors for COVID-19.

This study has several limitations. First, because the capacity for testing for COVID-19 patients varies among different countries, the reported COVID-19 cases may not fully represent the actual situation of COVID-19 outbreaks in some countries that could affect the accuracy of our predictive models. Second, the sample size is not sufficiently large in terms of the number of predictors we used. As a result, the *β*-coefficients of some variables were small, so that their associations with COVID-19 risk were ambiguous.

## Conclusions

Numerous factors may affect COVID-19 transmission and fatality, of which age, climate, social distancing, economic development, health investment, weight, and nutrient intake are most significant. Our comprehensive identification of the factors affecting COVID-19 transmission and fatality may provide new insights into the COVID-19 pandemic and advise effective strategies for preventing and migrating COVID-19 spread.

## Methods

### Materials

We defined 77 variables that could affect COVID-19 transmission and fatality in different countries. These variables included population density, life expectancy, sex ratio (number of males per 100 females), percentage of population aged < 10 years, percentage of population aged **≥** 70 years, percentage of urban population, percentages of population with body mass index (BMI) > 30 and BMI < 18, PM2.5 value (particles less than 2.5 micrometers in diameter), percentage of population using unimproved water sources, prevalence of tuberculosis, prevalence of diabetes, prevalence of HIV, smoking rate among people aged > 15 years, two immunization (DPT and measles) coverages among children ages 12-23 months, average age of childbirth, three types of geographic location (inland, island, and other), three temperature types (high (> 23.5 °C), medium ([0.7, 23.5] °C), low (< 0.7 °C)), humidity, latitude, longitude, five climate types (tropical, arid, temperate, cold, and other), four religions (Islam, Christianism, Buddhism, and other), governing system (unitary state or federation), Gini index, per capita income, universal health coverage index, government expenditure on health (% of GDP), hospital beds per 1,000 people, government expenditure on education (% of GDP), number of live audiences in major sports events,

number of participants in large-scale religious activities, number of participants in large-scale cultural events, birthrate, natimortality, prevalence of malnutrition, number of scientific papers published, human development index, international tourism expenditure, international tourism revenue, domestic tourism expenditure, air passenger traffic, railway length, vehicle usage, percentage of population using the internet, 16 classes of food and nutrient intake (vegetables, fruits, grain, dairy products, edible oil, fat, sugar, alcohol, trace elements, protein, vitamin A, B, C, D, E, and K), and four races (White, Black, Asian, and Arab). We downloaded the latest version of these data from major repositories of authoritative statistics. The major sports events, religious activities, and cultural events were the records from January to March 2020. A description of these variables is shown in Supplementary Table S1. Besides, we defined 55 variables that could affect COVID-19 transmission and fatality in different states of the United States. A description of these variables is shown in Supplementary Table S2. We collected confirmed COVID-19 cases, deaths, and case fatality rates (CFRs) in different countries and in the 50 U.S. states as of May 15, 2020, from the COVID-19 Dashboard of Johns Hopkins University ^1^. As suggested in a recent publication ^22^, we calculated the CFRs on a given day by dividing the number of COVID-19 deaths on that day by the number of cases 14 days before.

### Machine learning model

We defined the countries as having low or high COVID-19 cases, deaths, or CFRs using the 40^th^ and 60^th^ percentiles. We separated the data set into a training set and a test set, which contained three quarters and the remaining one quarter of the samples, respectively. We used the LASSO on the training set with 5-fold cross validation (CV) and selected the variables with a nonzero *β*-coefficient in the LASSO. Using the selected variables, we built a ridge logistic model on the training set and applied it to the test set. We estimated the sensitivity and specificity of the predictive model using the area under the receiver operating characteristic curve (AUC).

### Ranking variables by importance

For continuous variables, we discretized them using the top-down discretization method ^23^. We ranked the importance of all variables based on the chi-squared statistics. We performed data discretization using the R function “disc.Topdown” in the R package “discretization” and feature ranking using the R function “chi.squared” in the R package “mlr.”

### Statistical analysis

We performed comparisons of two groups using the one-sided Mann–Whitney U test. We evaluated the correlations of COVID-19 cases, deaths, and CFRs with other variables using the Spearman’s correlation test. The correlation test *P*-value and correlation coefficient (*ρ*) were reported.

## Data Availability

All data referred to in the manuscript are available.

## List of abbreviations

SARS-CoV-2: 2019 novel coronavirus
CFRs: case fatality rates
BMI: body mass index
*ρ*: correlation coefficient
AUC: the area under the receiver operating characteristic curve
CV: cross validation.

## Acknowledgments

Not applicable.

## Supplementary Materials

**Table S1.**
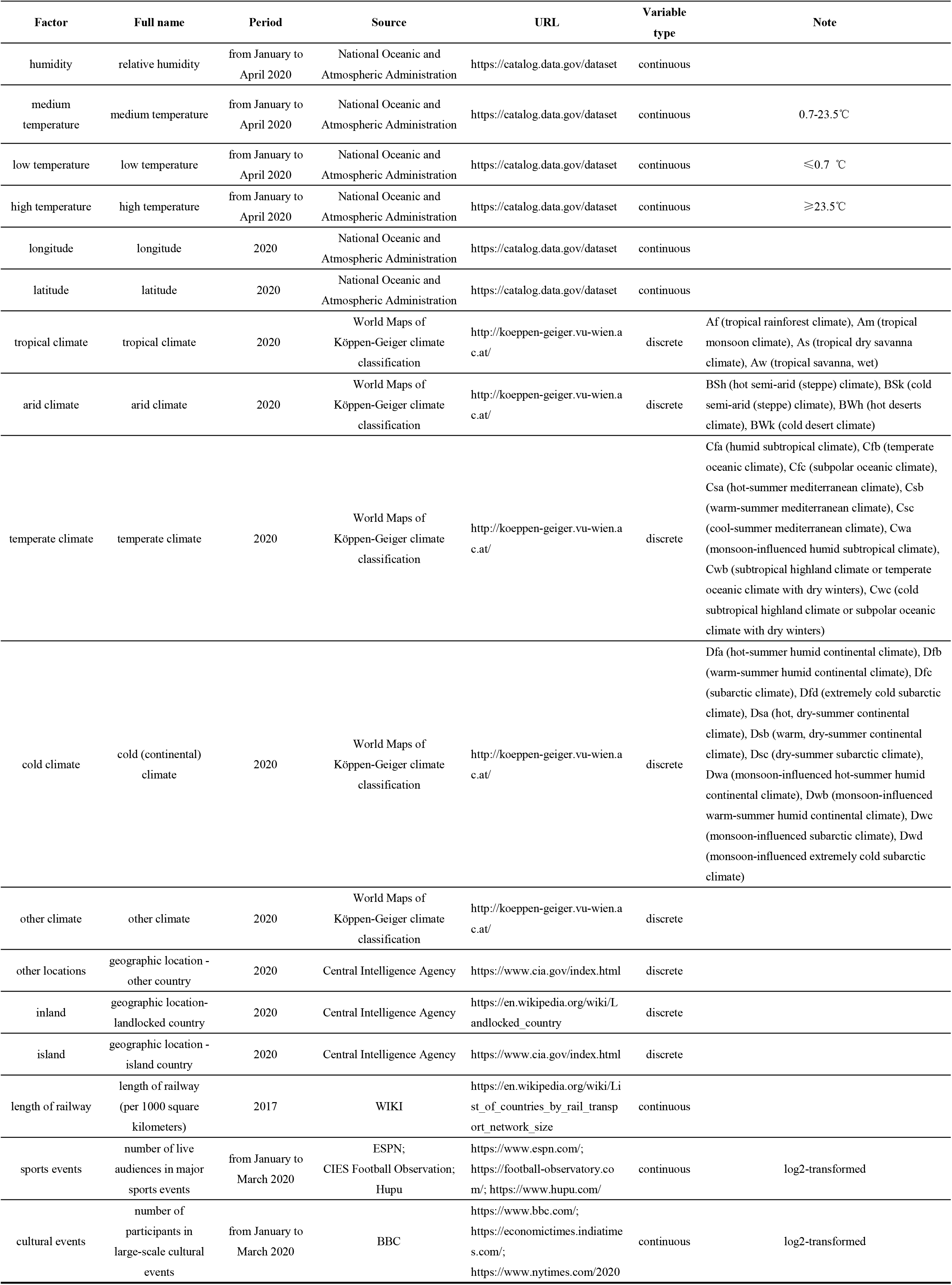
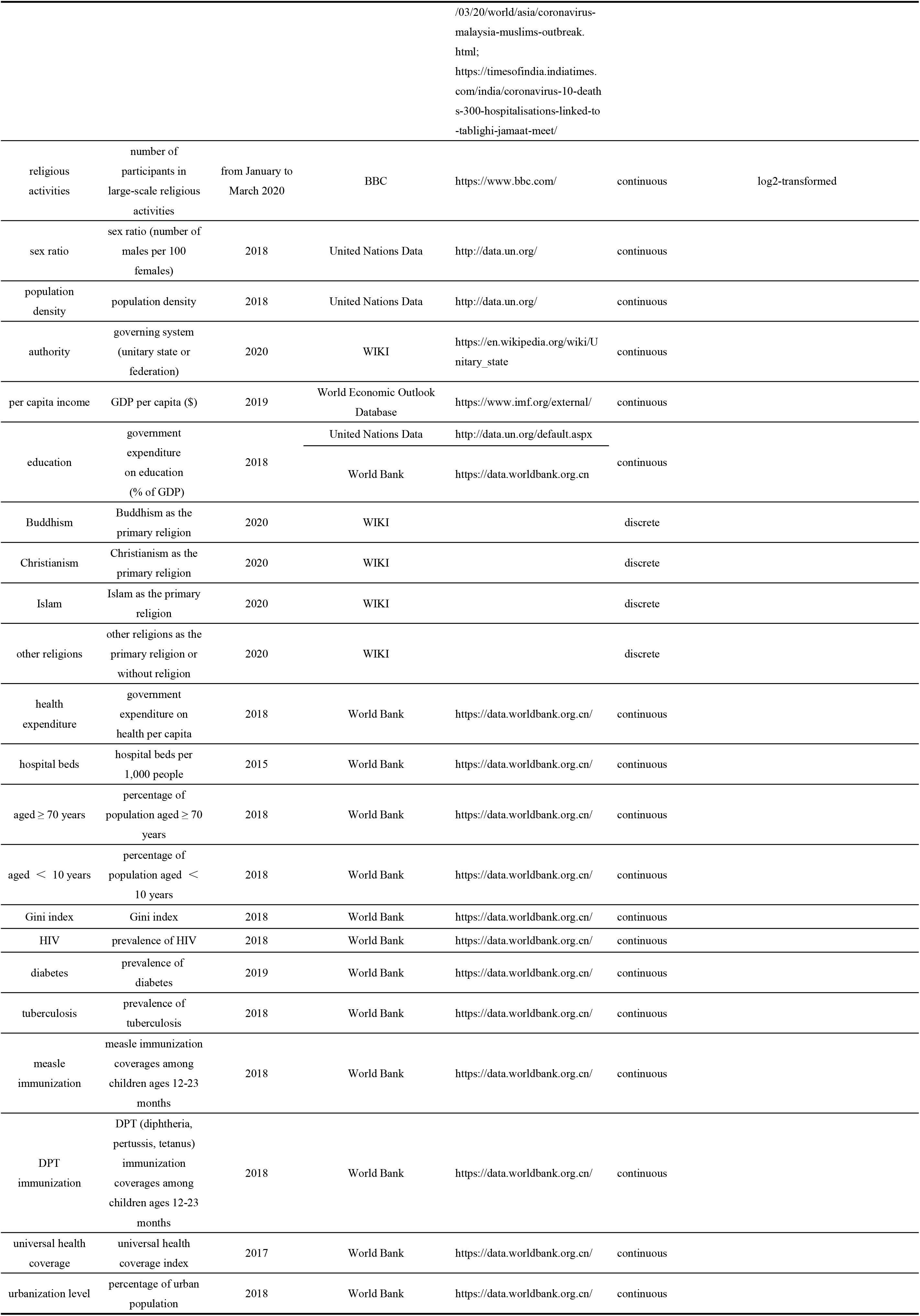
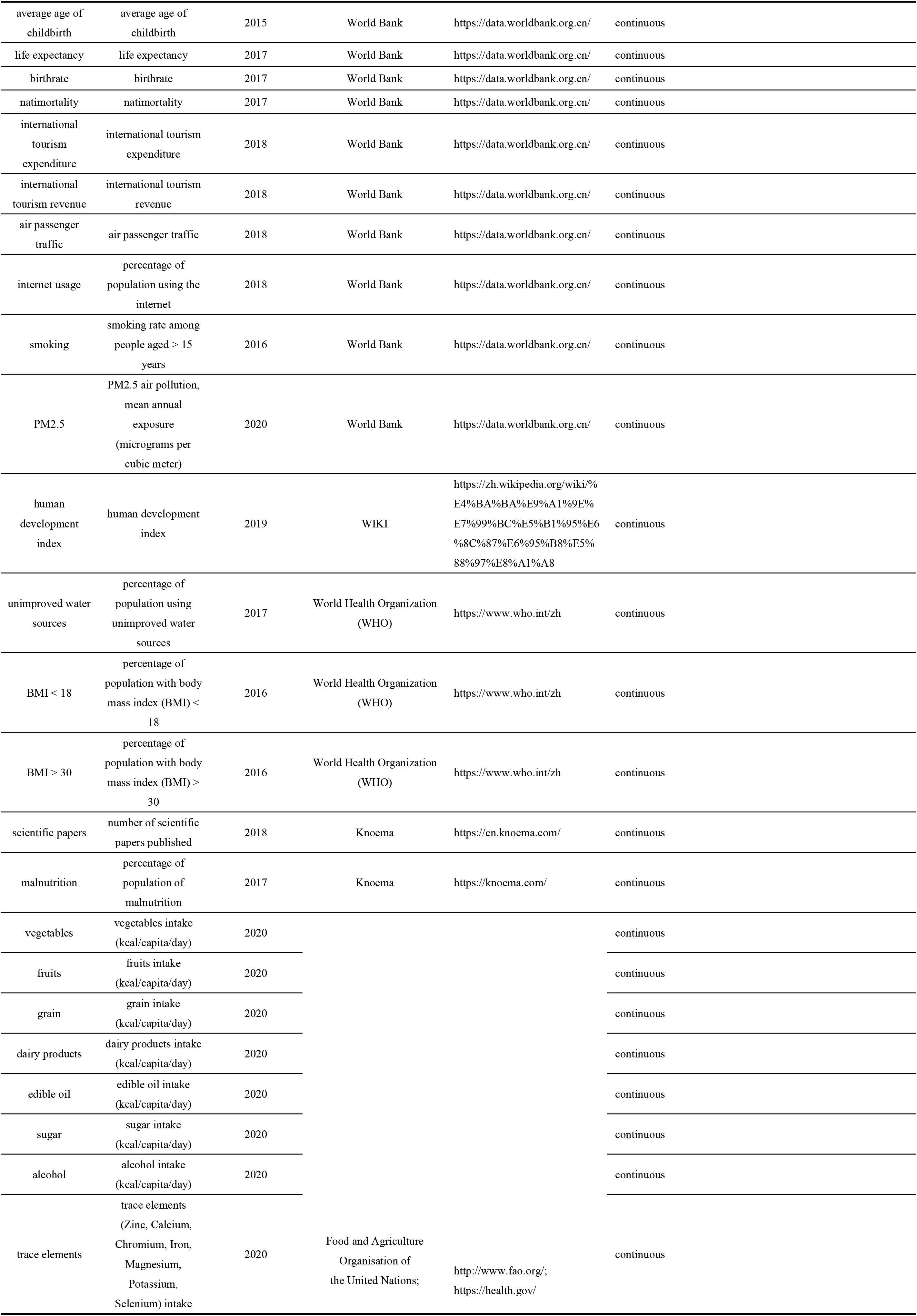
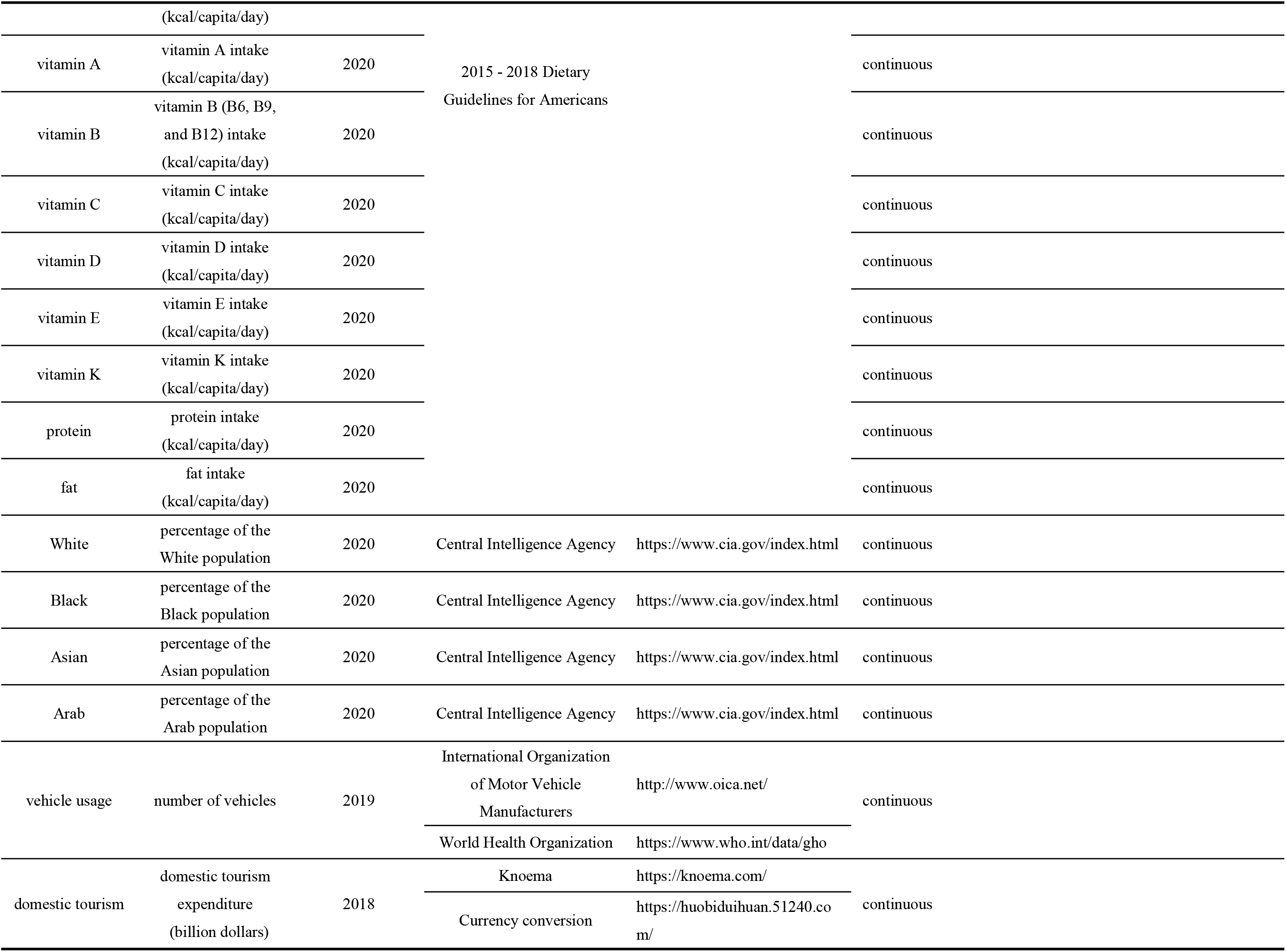
78 factors (variables) that could affect COVID-19 transmission and fatality in 154 countries.

|  |  |  |  |  |  |  |
| --- | --- | --- | --- | --- | --- | --- |
| Factor | Full name | Period | Source | URL | Variable type | Note |
| humidity | relative humidity | from January to April 2020 | National Oceanic and Atmospheric Administration | https://catalog.data.gov/dataset | continuous |  |
| medium temperature | medium temperature | from January to April 2020 | National Oceanic and Atmospheric Administration | https://catalog.data.gov/dataset | continuous | 0.7-23.5℃ |
| low temperature | low temperature | from January to April 2020 | National Oceanic and Atmospheric Administration | https://catalog.data.gov/dataset | continuous | ≤0.7 ℃ |
| high temperature | high temperature | from January to April 2020 | National Oceanic and Atmospheric Administration | https://catalog.data.gov/dataset | continuous | ≥23.5℃ |
| longitude | longitude | 2020 | National Oceanic and Atmospheric Administration | https://catalog.data.gov/dataset | continuous |  |
| latitude | latitude | 2020 | National Oceanic and Atmospheric Administration | https://catalog.data.gov/dataset | continuous |  |
| tropical climate | tropical climate | 2020 | World Maps of Köppen-Geiger climate classification | http://koeppen-geiger.vu-wien.ac.at/ | discrete | Af (tropical rainforest climate), Am (tropical monsoon climate), As (tropical dry savanna climate), Aw (tropical savanna, wet) |
| arid climate | arid climate | 2020 | World Maps of Köppen-Geiger climate classification | http://koeppen-geiger.vu-wien.ac.at/ | discrete | BSh (hot semi-arid (steppe) climate), BSk (cold semi-arid (steppe) climate), BWh (hot deserts climate), BWk (cold desert climate) |
| temperate climate | temperate climate | 2020 | World Maps of Köppen-Geiger climate classification | http://koeppen-geiger.vu-wien.ac.at/ | discrete | Cfa (humid subtropical climate), Cfb (temperate oceanic climate), Cfc (subpolar oceanic climate), Csa (hot-summer mediterranean climate), Csb (warm-summer mediterranean climate), Csc (cool-summer mediterranean climate), Cwa (monsoon-influenced humid subtropical climate), Cwb (subtropical highland climate or temperate oceanic climate with dry winters), Cwc (cold subtropical highland climate or subpolar oceanic climate with dry winters) |
| cold climate | cold (continental) climate | 2020 | World Maps of Köppen-Geiger climate classification | http://koeppen-geiger.vu-wien.ac.at/ | discrete | Dfa (hot-summer humid continental climate), Dfb (warm-summer humid continental climate), Dfc (subarctic climate), Dfd (extremely cold subarctic climate), Dsa (hot, dry-summer continental climate), Dsb (warm, dry-summer continental climate), Dsc (dry-summer subarctic climate), Dwa (monsoon-influenced hot-summer humid continental climate), Dwb (monsoon-influenced warm-summer humid continental climate), Dwc (monsoon-influenced subarctic climate), Dwd (monsoon-influenced extremely cold subarctic climate) |
| other climate | other climate | 2020 | World Maps of Köppen-Geiger climate classification | http://koeppen-geiger.vu-wien.ac.at/ | discrete |  |
| other locations | geographic location - other country | 2020 | Central Intelligence Agency | https://www.cia.gov/index.html | discrete |  |
| inland | geographic location-landlocked country | 2020 | Central Intelligence Agency | https://en.wikipedia.org/wiki/Landlocked_country | discrete |  |
| island | geographic location - island country | 2020 | Central Intelligence Agency | https://www.cia.gov/index.html | discrete |  |
| length of railway | length of railway (per 1000 square kilometers) | 2017 | WIKI | https://en.wikipedia.org/wiki/List_of_countries_by_rail_transport_network_size | continuous |  |
| sports events | number of live audiences in major sports events | from January to March 2020 | ESPN; CIES Football Observation; Hupu | https://www.espn.com/; https://football-observatory.com/; https://www.hupu.com/ | continuous | log2-transformed |
| cultural events | number of participants in large-scale cultural events | from January to March 2020 | BBC | https://www.bbc.com/; https://economictimes.indiatimes.com/; https://www.nytimes.com/2020 | continuous | log2-transformed |
|  |  |  |  | /03/20/world/asia/coronavirus-malaysia-muslims-outbreak.html;<br>https://timesofindia.indiatimes.com/india/coronavirus-10-deaths-300-hospitalisations-linked-to-tablighi-jamaat-meet/ |  |  |
| religious activities | number of participants in large-scale religious activities | from January to March 2020 | BBC | https://www.bbc.com/ | continuous | log2-transformed |
| sex ratio | sex ratio (number of males per 100 females) | 2018 | United Nations Data | http://data.un.org/ | continuous |  |
| population density | population density | 2018 | United Nations Data | http://data.un.org/ | continuous |  |
| authority | governing system (unitary state or federation) | 2020 | WIKI | https://en.wikipedia.org/wiki/Unitary_state | continuous |  |
| per capita income | GDP per capita (\$) | 2019 | World Economic Outlook Database | https://www.imf.org/external/ | continuous | |
| education | government expenditure on education (% of GDP) | 2018 | United Nations Data | http://data.un.org/default.aspx | continuous |  |
|  |  |  | World Bank | https://data.worldbank.org.cn |  |  |
| Buddhism | Buddhism as the primary religion | 2020 | WIKI |  | discrete |  |
| Christianism | Christianism as the primary religion | 2020 | WIKI |  | discrete |  |
| Islam | Islam as the primary religion | 2020 | WIKI |  | discrete |  |
| other religions | other religions as the primary religion or without religion | 2020 | WIKI |  | discrete |  |
| health expenditure | government expenditure on health per capita | 2018 | World Bank | https://data.worldbank.org.cn/ | continuous |  |
| hospital beds | hospital beds per 1,000 people | 2015 | World Bank | https://data.worldbank.org.cn/ | continuous |  |
| aged ≥ 70 years | percentage of population aged ≥ 70 years | 2018 | World Bank | https://data.worldbank.org.cn/ | continuous | |
| aged $<$ 10 years | percentage of population aged $<$ 10 years | 2018 | World Bank | https://data.worldbank.org.cn/ | continuous | |
| Gini index | Gini index | 2018 | World Bank | https://data.worldbank.org.cn/ | continuous |  |
| HIV | prevalence of HIV | 2018 | World Bank | https://data.worldbank.org.cn/ | continuous |  |
| diabetes | prevalence of diabetes | 2019 | World Bank | https://data.worldbank.org.cn/ | continuous |  |
| tuberculosis | prevalence of tuberculosis | 2018 | World Bank | https://data.worldbank.org.cn/ | continuous |  |
| measle immunization | measle immunization coverages among children ages 12-23 months | 2018 | World Bank | https://data.worldbank.org.cn/ | continuous |  |
| DPT immunization | DPT (diphtheria, pertussis, tetanus) immunization coverages among children ages 12-23 months | 2018 | World Bank | https://data.worldbank.org.cn/ | continuous |  |
| universal health coverage | universal health coverage index | 2017 | World Bank | https://data.worldbank.org.cn/ | continuous |  |
| urbanization level | percentage of urban population | 2018 | World Bank | https://data.worldbank.org.cn/ | continuous |  |

|  |  |  |  |  |  |
| --- | --- | --- | --- | --- | --- |
| average age of childbirth | average age of childbirth | 2015 | World Bank | https://data.worldbank.org.cn/ | continuous |
| life expectancy | life expectancy | 2017 | World Bank | https://data.worldbank.org.cn/ | continuous |
| birthrate | birthrate | 2017 | World Bank | https://data.worldbank.org.cn/ | continuous |
| natimortality | natimortality | 2017 | World Bank | https://data.worldbank.org.cn/ | continuous |
| international tourism expenditure | international tourism expenditure | 2018 | World Bank | https://data.worldbank.org.cn/ | continuous |
| international tourism revenue | international tourism revenue | 2018 | World Bank | https://data.worldbank.org.cn/ | continuous |
| air passenger traffic | air passenger traffic | 2018 | World Bank | https://data.worldbank.org.cn/ | continuous |
| internet usage | percentage of population using the internet | 2018 | World Bank | https://data.worldbank.org.cn/ | continuous |
| smoking | smoking rate among people aged > 15 years | 2016 | World Bank | https://data.worldbank.org.cn/ | continuous |
| PM2.5 | PM2.5 air pollution, mean annual exposure (micrograms per cubic meter) | 2020 | World Bank | https://data.worldbank.org.cn/ | continuous |
| human development index | human development index | 2019 | WIKI | https://zh.wikipedia.org/wiki/%E4%BA%BA%E9%A1%9E%E7%99%BC%E5%B1%95%E6%8C%87%E6%95%B8%E5%88%97%E8%A1%A8 | continuous |
| unimproved water sources | percentage of population using unimproved water sources | 2017 | World Health Organization (WHO) | https://www.who.int/zh | continuous |
| BMI < 18 | percentage of population with body mass index (BMI) < 18 | 2016 | World Health Organization (WHO) | https://www.who.int/zh | continuous |
| BMI > 30 | percentage of population with body mass index (BMI) > 30 | 2016 | World Health Organization (WHO) | https://www.who.int/zh | continuous |
| scientific papers | number of scientific papers published | 2018 | Knoema | https://cn.knoema.com/ | continuous |
| malnutrition | percentage of population of malnutrition | 2017 | Knoema | https://knoema.com/ | continuous |
| vegetables | vegetables intake (kcal/capita/day) | 2020 | Food and Agriculture Organisation of the United Nations; | http://www.fao.org/;<br>https://health.gov/ | continuous |
| fruits | fruits intake (kcal/capita/day) | 2020 |  |  | continuous |
| grain | grain intake (kcal/capita/day) | 2020 |  |  | continuous |
| dairy products | dairy products intake (kcal/capita/day) | 2020 |  |  | continuous |
| edible oil | edible oil intake (kcal/capita/day) | 2020 |  |  | continuous |
| sugar | sugar intake (kcal/capita/day) | 2020 |  |  | continuous |
| alcohol | alcohol intake (kcal/capita/day) | 2020 |  |  | continuous |
| trace elements | trace elements (Zinc, Calcium, Chromium, Iron, Magnesium, Potassium, Selenium) intake (kcal/capita/day) | 2020 | Food and Agriculture Organisation of the United Nations; | http://www.fao.org/;<br>https://health.gov/ | continuous |
| vitamin A | vitamin A intake<br>(kcal/capita/day) | 2020 | 2015 - 2018 Dietary<br>Guidelines for Americans |  | continuous |
| vitamin B | vitamin B (B6, B9,<br>and B12) intake<br>(kcal/capita/day) | 2020 |  |  | continuous |
| vitamin C | vitamin C intake<br>(kcal/capita/day) | 2020 |  |  | continuous |
| vitamin D | vitamin D intake<br>(kcal/capita/day) | 2020 |  |  | continuous |
| vitamin E | vitamin E intake<br>(kcal/capita/day) | 2020 |  |  | continuous |
| vitamin K | vitamin K intake<br>(kcal/capita/day) | 2020 |  |  | continuous |
| protein | protein intake<br>(kcal/capita/day) | 2020 |  |  | continuous |
| fat | fat intake<br>(kcal/capita/day) | 2020 |  |  | continuous |
| White | percentage of the<br>White population | 2020 | Central Intelligence Agency | https://www.cia.gov/index.html | continuous |
| Black | percentage of the<br>Black population | 2020 | Central Intelligence Agency | https://www.cia.gov/index.html | continuous |
| Asian | percentage of the<br>Asian population | 2020 | Central Intelligence Agency | https://www.cia.gov/index.html | continuous |
| Arab | percentage of the<br>Arab population | 2020 | Central Intelligence Agency | https://www.cia.gov/index.html | continuous |
| vehicle usage | number of vehicles | 2019 | International Organization<br>of Motor Vehicle<br>Manufacturers | http://www.oica.net/ | continuous |
|  |  |  | World Health Organization | https://www.who.int/data/gho |  |
| domestic tourism | domestic tourism<br>expenditure<br>(billion dollars) | 2018 | Knoema | https://knoema.com/ | continuous |
|  |  |  | Currency conversion | https://huobiduihuan.51240.com/ |  |

**Table S2.**
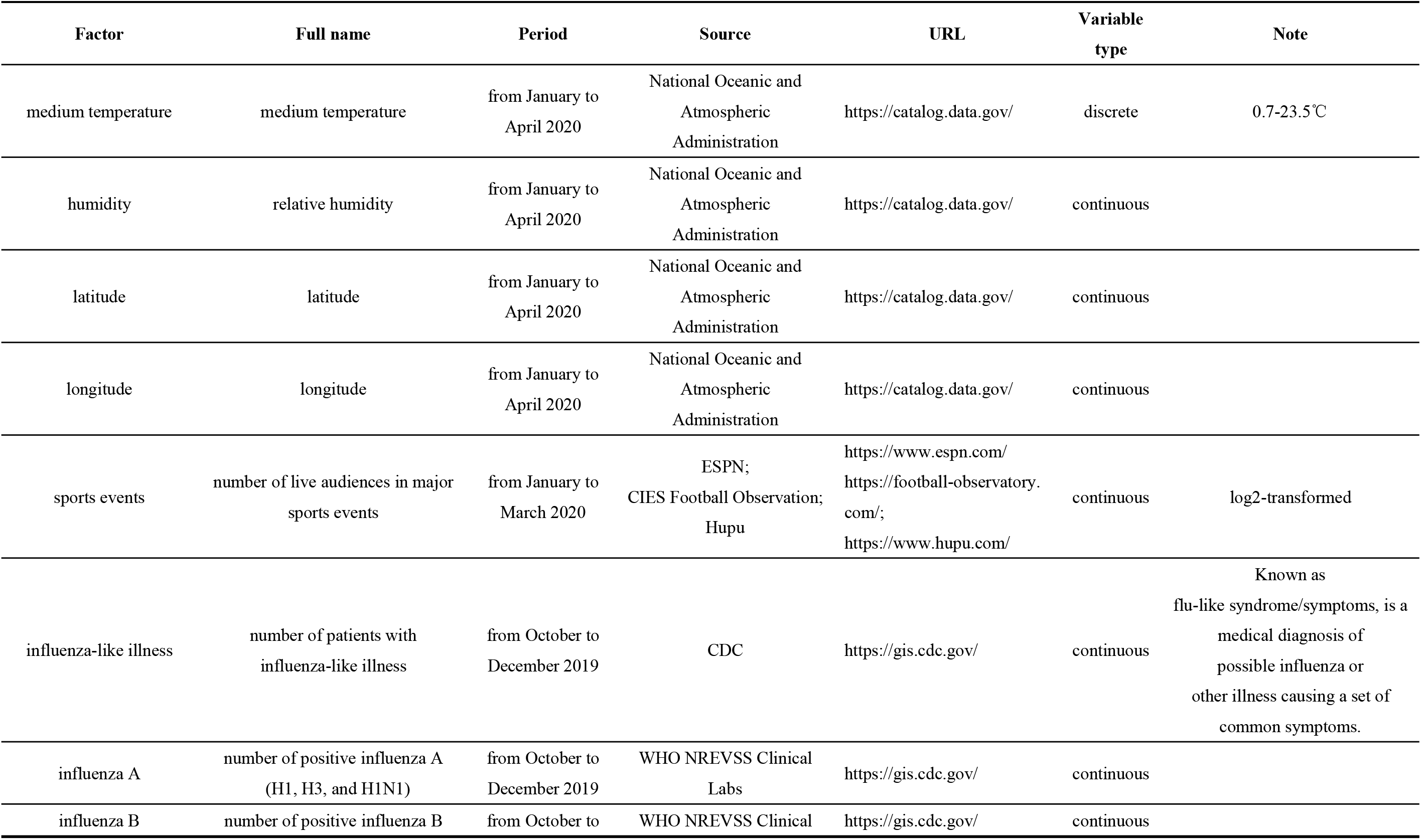
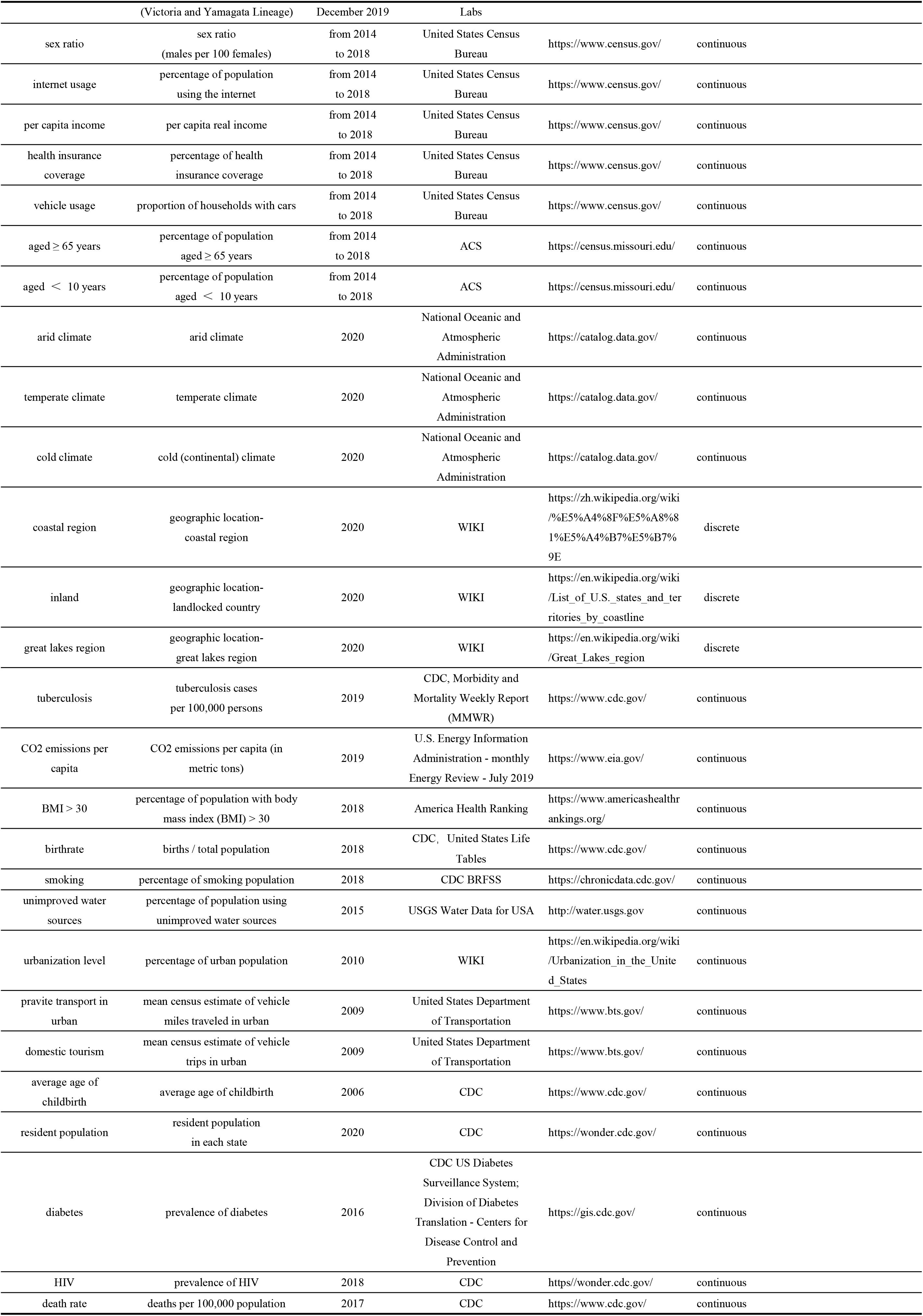
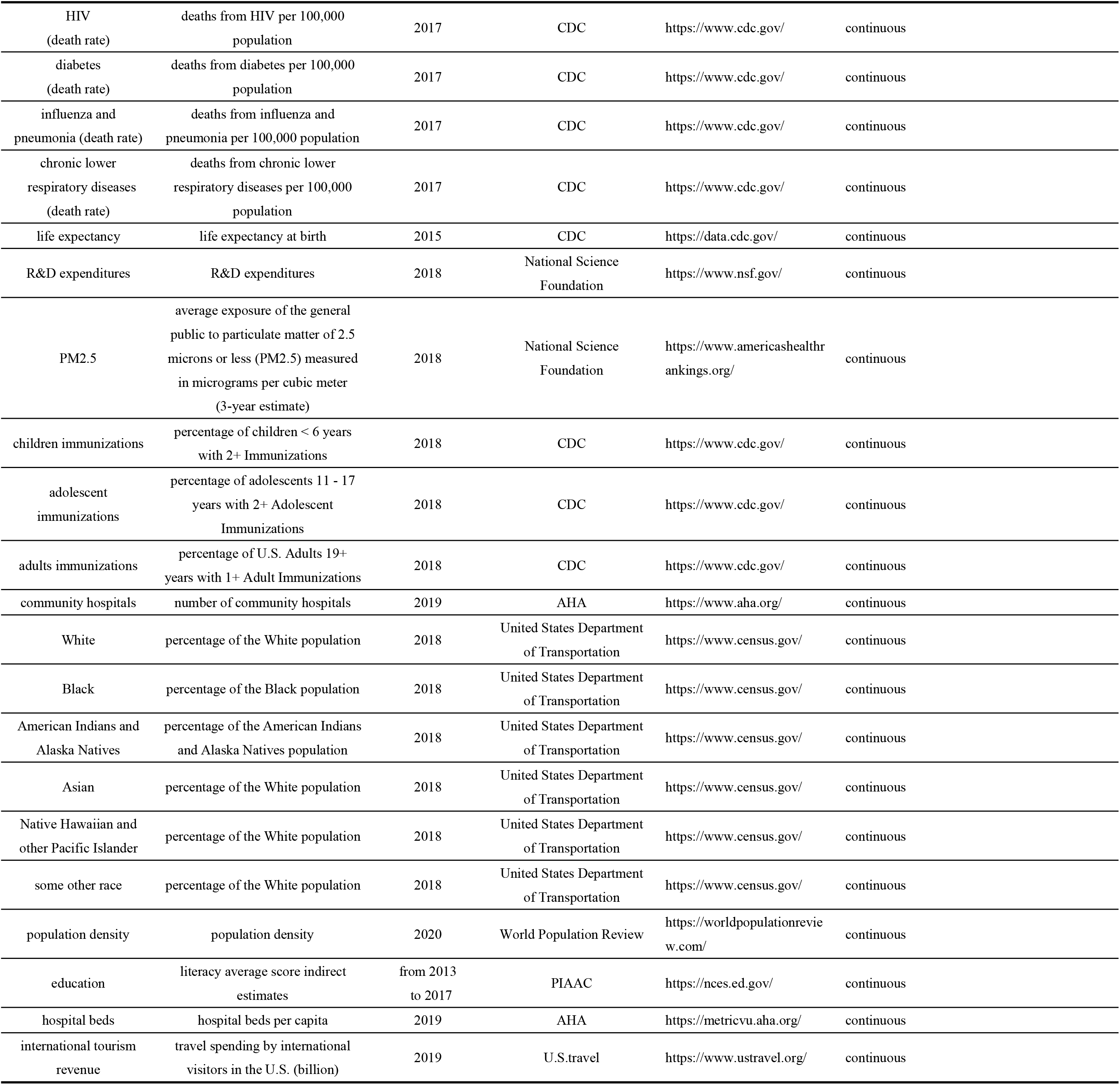
55 factors (variables) that could affect COVID-19 transmission and fatality in the 50 U.S. states.

